# Interleukin-6 predicts depersonalisation and C-reactive protein predicts derealisation: A longitudinal study using the Avon Longitudinal Study of Parents and Children dataset

**DOI:** 10.1101/2025.08.26.25334112

**Authors:** Evelyn Dilkes, Katie Daughters, Helge Gillmeister

## Abstract

**Background:** Depersonalisation (DP) and derealisation (DR) are dissociative symptoms that impair perception of one’s self and environment. There is limited understanding of the biological mechanisms underpinning persistent DPDR, but emerging evidence suggests the involvement of inflammatory processes. This study investigates the longitudinal association between inflammatory markers (interleukin-6 [IL-6] and C-reactive protein [CRP]) and DPDR symptoms in a population-based cohort.

**Methods:** We used data from the Avon Longitudinal Study of Parents and Children. IL-6 was measured at age 9 years; CRP at ages 9, 15, and 24 years. DPDR were assessed at ages 12, 17, and 24 years. Generalised linear mixed models analysed relationships, adjusting for sex, ethnicity, and social position.

**Results:** Higher IL-6 at age 9 years significantly predicted increased odds of DP at age 24 (aOR = 1.52, p = 0.004) in comparison to DP at age 12. Higher CRP levels at age 9 years were associated with increased odds of DR at age 12 (aOR = 1.43, p = 0.010). However, a significant negative association was observed between CRP and DR at age 24 (aOR = 0.45, p < 0.001) in comparison to DR at age 12. IL-6 and CRP were not associated with DR and DP, respectively, at any age.

**Conclusions:** Findings suggest distinct inflammatory profiles for DP and DR. Childhood IL-6 elevation predicts later DP, while CRP shows an age-dependent association with DR. This may reflect allostatic load or immune adaptation. Future studies should explore neuroinflammatory pathways, adversity, and intervention potential using dimensional measures of DPDR.

## Introduction

Depersonalisation (DP) and derealisation (DR) are dissociative symptoms characterised by profound alterations in self-perception and perception of the external world. DP involves feelings of detachment from one’s thoughts, emotions, body, or actions, often accompanied by perceptual distortions, a disrupted sense of time, emotional numbing, and a diminished sense of self (American Psychiatric Association, 2013). DR, in contrast, refers to an altered perception of the external environment, where surroundings may appear unreal, dreamlike, or visually distorted (American Psychiatric Association, 2013). These symptoms can be adaptive and frequently manifest in response to traumatic stress, but can become persistent and therefore dysfunctional (Simeon et al., 2004). If symptoms do not remit, individuals may be diagnosed with Derealisation-Depersonalisation Disorder (DDD), which is considered ‘uncommon’ (approximately 1-2% of the population may qualify for a diagnosis; Hunter, Sierra & David, 2004; Yang et al., 2022), However, given that up to 74% of people experience such symptoms at some point in their lives (Hunter, Sierra, & David, 2004), these diagnosis rates likely underestimate the true scale of the problem. Attaining a diagnosis of DDD can take between 7-12 years (Baker et al., 2003; Michal et al., 2016). This delay is compounded by a general lack of awareness among healthcare providers, leaving individuals with DDD vulnerable to misdiagnosis or unproductive medical consultations (Medford et al., 2005). Therefore, there is an increased likelihood that many individuals with DPDR remain undiagnosed, and larger population-based studies are necessary to better capture the full spectrum of DPDR experiences.

DDD has been associated with dysregulation of the limbic system (Sierra & Berrios, 1998; Phillips et al., 2001), autonomic nervous system (Millman et al., 2024), and hypothalamic-pituitary-adrenal (HPA) axis (Simeon et al., 2001; Simeon et al., 2007). More recent research, however, has begun to investigate the role of inflammatory in dissociative symptoms, including DPDR. Elevated inflammatory markers, such as C-reactive protein (CRP) and tumor necrosis factor-alpha (TNF-α), have been associated with dissociative symptoms in various psychiatric conditions (Powers et al., 2019; Bizik et al., 2011). Additionally, individuals with dissociative PTSD subtypes exhibit increased CRP levels compared to those with non-dissociative PTSD, suggesting a link between inflammation and dissociative processes (Jarkas et al., 2021). Furthermore, people with chronic inflammatory conditions may be at increased risk of mental health issues (Rakshasa-Loots et al., 2025), including dissociation. For example, inflammatory bowel disease, are considerably more likely to exhibit dissociative symptoms compared to healthy individuals (Ferrarese et al., 2021).

Interleukin-6 (IL-6) and CRP are of interest in dissociation research as they are biological markers that can offer insight into the biological embedding of early adversity (Berens et al., 2017). For example, lack of home ownership or low parental education, low social position and self-reported childhood trauma, are significantly associated, and can predict, increased concentrations of IL-6 and CRP (Miller & Chen., 2007; Packard et al., 2011; Carpenter et al., 2010; Lin et al., 2016). Similarly, early life adversity is highly associated with dissociation both independently (Thomson & Jaque, 2018; Vonderlin et al., 2018; King et al., 2020; Quiñones, 2022) and as a transdiagnostic symptom (for review see Rafiq, Campodonico & Varese, 2018) and therefore, CRP and IL-6 are likely candidates to play an inflammatory role in dissociative symptoms.

Only one study has investigated inflammation and DDD specifically, identifying dysregulated inflammatory markers, including reduced CRP and complement C1q subcomponent B, alongside increased alpha-1-antichymotrypsin in people with DDD (Zheng et al., 2024). Although based on only a small clinical sample, this study points to potential chronic low-grade inflammation in people with DDD.

Evidence of the role of IL-6 is mixed: one study found increased levels of IL-6 were strongly correlated with somatoform dissociation (involving disturbances or alterations in motor and sensory functioning, perception, or bodily experience, but without a known medical cause), but not with a broader measure of dissociation (Bob et al., 2010), in a sample of 40 inpatients with unipolar depression.

Despite growing insights into inflammatory mechanisms underlying DPDR, research remains limited by small, clinical sample sizes. Such studies, focused on diagnosed patients, restrict statistical power and generalisability to the broader population experiencing sub-clinical or undiagnosed DPDR. Expanding sample sizes and incorporating diverse cohorts are crucial to improve findings’ robustness and advance understanding of treatment targets. The current study aimed to address these limitations by investigating the effect of elevated inflammatory markers on DPDR symptoms in a large, prospective general population cohort, thereby providing novel insights into the biological mechanisms of DDD.

To the best of our knowledge, the only UK dataset to hold data on both DPDR and inflammatory markers is the Avon Longitudinal Study of Parents and Children (ALSPAC), containing over three decades of data from children born in the early 1990s. Previous research has effectively utilised the ALSPAC dataset to investigate the impact of IL-6 and CRP on depression (Khandaker et al., 2014; Osimo et al., 2020); psychosis (Perry et al., 2019; 2021; Donnelly et al., 2022) and anxiety (Mongan et al., 2023; Khandaker et al., 2016). Therefore, the ALSPAC dataset is a reliable source of inflammation data that is well poised for testing the longitudinal impact of inflammatory markers on DPDR.

### Study Hypotheses

This study examined whether elevated IL-6 and CRP levels predicted later symptoms of DP and DR, adjusting for key demographics. IL-6, measured at age 9, was used to assess long-term associations with DP and DR at ages 12, 17, and 24. CRP, measured at ages 9, 15, and 24, enabled analysis of its shorter-term effects across the same time points. DP and DR were analysed separately in line with evidence suggesting distinct neurobiological underpinnings (Murphy, 2023).

Hypotheses 1 and 2: Higher IL-6 levels at age 9 years will predict increased odds of experiencing depersonalisation symptoms (H1) and derealisation symptoms (H2) at ages 12, 17, and 24 years, after adjusting for sex, ethnicity, and social position.

Hypotheses 3 and 4: Higher CRP levels at each developmental stage (ages 9, 15, and 24 years) will predict increased odds of experiencing depersonalisation symptoms (H3) and derealisation symptoms (H4) at ages 12, 17, and 24 years, after adjusting for sex, ethnicity, and social position.

## Methods and Materials

### Cohort Description and Sample Sizes

This study used child data from ALSPAC (Harris et al., 2009; Boyd et al., 2013; Fraser et al., 2013; Northstone et al., 2019). This study used data from the Avon Longitudinal Study of Parents and Children (ALSPAC), a UK-based birth cohort designed to investigate factors influencing health, development, and disease from early life onwards. The cohort initially enrolled 14,541 pregnancies. A comprehensive overview of available data can be found on the ALSPAC website (http://www.bristol.ac.uk/alspac/researchers/our-data/). Ethical approval was granted by the ALSPAC Law and Ethics Committee and local research ethics committees. Informed consent was obtained in accordance with ethical guidelines in place at the time. For full details on the use of the ALSPAC cohort in this study, please see Supplementary Information, Section 1. Study data were collected and managed using REDCap electronic data capture tools hosted at the University of Bristol (Harris et al., 2009) REDCap (Research Electronic Data Capture) is a secure, web-based software platform designed to support data capture for research studies.

The present study focused on the inflammatory markers IL-6 and CRP, both of which were log-transformed prior to analysis. IL-6 was measured at age 9, and CRP at ages 9, 15, and 24 (see full details below). DP and DR symptoms were assessed at ages 12, 17, and 24. Participants reporting acute infections at the time of blood collection or within the preceding week, or with CRP values ≥10 mg/L, were excluded. Final sample sizes following these exclusions are presented in Table 1.

**Table 1:** Demographic information for participants at baseline (age 12)

| Demographic | CRP Dataset | IL-6 Dataset |
| --- | --- | --- |
|  | N (%) |  |
| **Total** | 3,774 | 2,388 |
| **Sex** |  |  |
| Male | 1891 (50) | 1135 (47) |
| Female | 1883 (50) | 1253 (53) |
| **Ethnicity** |  |  |
| White | 3462 (92) | 2173 (91) |
| Non-white | 131 (3) | 92 (4) |
| Missing | 217 (5) | 123 (5) |

| **Registrar General's Social Class Scale - Father's employment** |  |  |
| --- | --- | --- |
| I (Professional) | 418 (11) | 321 (13) |
| II (Managerial / Technical) | 1253 (33) | 804 (34) |
| III – N (Skilled non-manual occupations) | 434 (11) | 286 (12) |
| III – M (Skilled manual occupations) | 900 (24) | 556 (23) |
| IV – Partly-skilled occupations | 265 (7) | 132 (6) |
| V – Unskilled occupations | 66 (2) | 40 (2) |
| Missing | 438 (11) | 249 (10) |

### Inflammatory Marker Assessment

Blood samples were collected from non-fasting participants, immediately processed, and stored at −80°C. Inflammatory markers were assayed in 2008, ensuring samples were preserved without prior freeze-thaw cycles. IL-6 was measured using enzyme-linked immunosorbent assay. CRP was measured using an automated particle-enhanced immunoturbidimetric assay. All interassay coefficients of variation were less than 5%, ensuring high reliability.

### Depersonalisation and Derealisation Assessment

DP and DR were assessed at ages 12, 17 and 24. For DP, individuals were asked if they had “ever felt that they were not a real person, not part of the living world”, and for DR, individuals were asked if they had “ever felt that the world was unreal, that things around them were like a stage set”. Participants responded on a 3-point scale from ‘never’ to ‘frequently’. Participants reporting ‘frequently’ or ‘sometimes’ experiencing DP or DR were coded as 1 (experienced), while those who had never experienced these symptoms were coded as 0, creating a dichotomous variable for each DP and DR.

### Secondary Variables

To account for factors known to influence inflammation, we adjusted for child ethnicity, sex, and social position (Casimir et al., 2010; Schmeer & Tarrence, 2018). Ethnicity was based on the mother’s report of her own and her partner’s ethnic group during pregnancy, classified as white or non-white (non-white if either parent identified as such). Sex at birth was recorded as male or female. Social position was based on paternal occupation during pregnancy, classified using the UK Registrar General’s Social Class system (Szreter, 1984) as reported in the ALSPAC dataset.

## Data Analysis

This study was preregistered on AsPredicted (registration number 142,136). In line with ALSPAC’s governance protocols, raw data cannot be publicly shared; access is granted through a formal application process (https://www.bristol.ac.uk/alspac/). Data were analysed in RStudio (v3.3.4) using the dplyr (Wickham et al., 2023) and glmmTMB (Brooks et al., 2017) packages.

Generalized Linear Mixed Models (GLMMs) with a binomial distribution and logit link were used to account for repeated measures and the binary nature of DPDR outcomes. Random intercepts were included for participant ID. IL-6 (age 9) and CRP (ages 9, 15, 24) were primary predictors, with sex, ethnicity, and social position as covariates. Separate datasets were used to maximise power due to differing data availability (see Cohort Description).

For predictors that did not vary over time (IL-6 and CRP at age 9), additional cross-sectional GLMs were run to examine baseline associations with DPDR at age 12. CRP at age 9 was also modelled longitudinally for consistency (see Supplementary Information, Section 2). GLMMs used maximum likelihood estimation and assumed data were missing at random.

## Results

GLMM findings for associations between inflammatory markers and symptoms of DP and DR can be seen in Tables 2 (IL-6) and 3 (CRP) below.

### Interleukin-6

Log-transformed IL-6 at 9 years was not significantly associated with DP at 12 years (aOR = 01.05, 95% CI: 0.94 - 1.15, p = .374). Further, in relation to DP scores at age 12, log-transformed IL-6 did not significantly predict increased odds of experiencing DP at age 17 (aOR = 0.95, 95% CI: 0.66 – 1.38, p = 0.800). A significant association was found between higher log-transformed IL-6 levels at age 9 and an increased likelihood of DP symptoms at age 24 (aOR = 1.52, 95% CI: 1.14 – 2.03, p = 0.004). This indicates that for every one-unit increase in log-transformed IL-6, the odds of experiencing DP symptoms increase by 52% relative to the odds at age 12, thereby partially supporting Hypothesis 1. There was no support for Hypothesis 2 as there were no significant relationships between log-transformed IL-6 and DR symptoms at any time point.

**Table 2:**
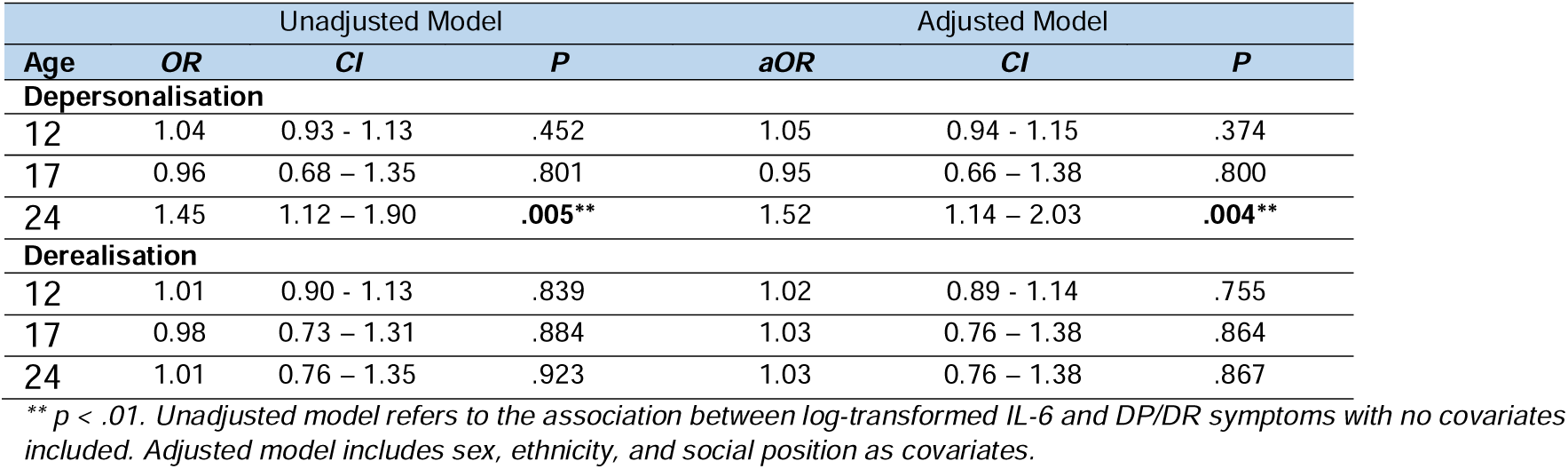
Unadjusted and adjusted OR for the association between log-transformed IL-6 at age 9 and DP and DR at ages 12, 17, and 24.

| Age | Unadjusted Model |  |  | Adjusted Model |  |  |
| --- | --- | --- | --- | --- | --- | --- |
|  | OR | CI | P | aOR | CI | P |
| **Depersonalisation** |  |  |  |  |  |  |
| 12 | 1.04 | 0.93 - 1.13 | .452 | 1.05 | 0.94 - 1.15 | .374 |
| 17 | 0.96 | 0.68 – 1.35 | .801 | 0.95 | 0.66 – 1.38 | .800 |
| 24 | 1.45 | 1.12 – 1.90 | **.005\*\*** | 1.52 | 1.14 – 2.03 | **.004\*\*** |
| **Derealisation** |  |  |  |  |  |  |
| 12 | 1.01 | 0.90 - 1.13 | .839 | 1.02 | 0.89 - 1.14 | .755 |
| 17 | 0.98 | 0.73 – 1.31 | .884 | 1.03 | 0.76 – 1.38 | .864 |
| 24 | 1.01 | 0.76 – 1.35 | .923 | 1.03 | 0.76 – 1.38 | .867 |

\*\* $p < .01$ . Unadjusted model refers to the association between log-transformed IL-6 and DP/DR symptoms with no covariates included. Adjusted model includes sex, ethnicity, and social position as covariates.

### C-reactive Protein

A significant positive association was observed between log-transformed CRP at age 9 and DR at age 12 years (aOR = 1.43, 95% CI: 1.09 – 1.90, p = 0.010). This indicates that for every one-unit increase in log-transformed CRP, the odds of experiencing DR increase by 43%. Further, the results indicated that, in relation to DR at age 12, there was no significant relationship between log-transformed CRP at age 15 and DR at age 17 years, suggesting that CRP at age 15 did not significantly predict increased or decreased prevalence of DR at age 17 (aOR = 0.74, 95% CI:0.45 – 1.21, p = 0.233). However, a significant negative association was found between log-transformed CRP at age 24 and DR at age 24 (aOR = 0.45, 95% CI: 0.27– 0.72, p < 0.001). This indicates that for every one-unit increase in log-transformed CRP, the odds of experiencing DR decrease by 55%. These findings provide partial support for hypothesis 4. There was no support for Hypothesis 3 as there were no significant relationships between CRP and DP symptoms at any time point.

**Table 3:** Unadjusted and adjusted OR for the association between log-transformed CRP at ages 9, 15 and 24 and DP and DR at ages 12, 17, and 24.

| Age | Unadjusted Model |  |  | Adjusted Model |  |  |
| --- | --- | --- | --- | --- | --- | --- |
|  | OR | CI | P | aOR | CI | P |
| **Depersonalisation** |  |  |  |  |  |  |
| 12 | 1.00 | 0.77 – 1.31 | .974 | 0.99 | 0.74 – 1.32 | .959 |
| 17 | 0.89 | 0.51 – 1.57 | .688 | 1.02 | 0.55 – 1.88 | .960 |
| 24 | 0.85 | 0.49 – 1.46 | .554 | 0.83 | 0.46 – 1.51 | .536 |
| **Derealisation** |  |  |  |  |  |  |
| 12 | 1.45 | 1.13 – 1.88 | .004** | 1.43 | 1.09 – 1.90 | .010* |
| 17 | 0.78 | 0.50 – 1.22 | .277 | 0.74 | 0.45 – 1.21 | .233 |
| 24 | 0.51 | 0.32 – 0.79 | .002** | 0.45 | 0.27 – 0.72 | <.001*** |

\*\*\* $p < .001$ , \*\* $p < .01$ , \* $p < .05$ . Unadjusted model refers to the association between log-transformed CRP and DP/DR symptoms with no covariates included. Adjusted model includes sex, ethnicity, and social position as covariates.

## Discussion

This is the first longitudinal study to investigate the role of inflammatory markers in DPDR symptoms in a large, general population sample, providing novel insights into the maintenance of these symptoms over time. The findings were mixed, with two hypotheses partially supported and two rejected. Early-life IL-6 elevation predicted a 52% increase in the odds of experiencing DP symptoms at age 24 years, suggesting a delayed effect of childhood inflammation. This effect was specific to DP, as there were no significant association between IL-6 and DR symptoms at any age. In contrast, CRP levels were not associated with DP symptoms at any age. Instead, CRP at age 9 was associated with a 43% increased probability of DR at age 12 years. A few years later, CRP at age 15 was no longer associated with DR at age 17 years. In adulthood (age 24 years), CRP was associated with a 55% *lower* probability of experiencing DR.

Our study is the first to observe the longitudinal effect of both IL-6 and CRP on sub-clinical symptoms of DDD, and the first to investigate the effect of these inflammatory markers on DP and DR separately. Our findings contribute to a growing body of literature suggesting that immune system dysregulation may influence dissociative symptoms (Power et al., 2019; Bizik et al., 2011, Jarkas et al., 2021; Bob et al., 2010) and DPDR more specifically (Zheng et al., 2024). Identification that increased levels of IL-6 at 9 years was predictive of DP at 24 years of age suggest that early life low-grade inflammation has an impact into adulthood. This is in line with previous studies that identify such a relationship between IL-6 and other mental health outcomes, such as depression (Chu et al., 2019), hypomania (Hayes et al., 2016) and depressive and psychotic symptoms in young adulthood (Perry et al., 2021). The present study’s use of a binary DP measurement prevented assessing a dose-response relationship, which should be a key area for future research.

The only prior study to investigate the relationship between inflammatory markers and DP and DR separately found lower CRP levels in 30 individuals with DDD and 32 healthy controls, matched for sex and age (average age 24 years, 7:23 gender ratio for females and males, Chinese Han ethnicity; Zheng et al., 2024). In that study, CRP downregulation was significantly correlated with ‘emotional numbing’ (reflective of DP) but not with ‘unreality of surroundings’ (reflective of DR). These findings contrast with the current study, where lower CRP levels at a similar age (24 years) was associated with ‘feeling that the world was unreal’ (reflective of DR) but not with ‘feeling that they were not a real person’ (reflective of DP). This may be due to differing ethnicity, clinical severity or time point of investigation (2015/16 vs. 2023). Nevertheless, both studies suggest a relationship between lower CRP and a dimension of DPDR in early adulthood, although the specific dimension (DP vs. DR) remains unclear and requires further research.

The present study found a reversal in the impact of CRP on DR, with increased CRP predicting DR at baseline (age 12) but lower CRP predicting DR by age 24 years. This may seem counterintuitive given previous research showing increased CRP is associated with increased psychiatric symptoms (Goldsmith et al., 2023) and increased inflammatory markers being more broadly associated with dissociative symptoms (Power et al., 2019; Bizik et al., 2011; Jarkas et al., 2021; Bob et al., 2010). However, the pattern we observed could reflect immunosuppression from chronic stress (Dhabhar, 2014) or immune habituation to stressors over time (Barthel et al., 2025). Suppression or habituation might explain why CRP decreases while DR symptoms persist, suggesting that the psychological impact of prior stressors may endure after the body’s acute inflammatory response has subsided.

Additionally, the persistence of DR symptoms despite reduced CRP levels at age 24, may reflect a shift from external psychosocial stressors (e.g., ACEs) to internal stressors, such as catastrophic cognitions and hypervigilance related to the DPDR symptoms themselves. This aligns with the cognitive-behavioural model of DPDR (Hunter, Salkovskis & David, 2014), which posits that dissociative symptoms, initially an adaptive response to stress, can become self-sustaining through psychological mechanisms like catastrophic interpretations and hypervigilance. These cognitive factors can maintain DPDR even after the initial stressor or biological inflammatory response has subsided. Notably, such internal or cognitive stressors may be associated with different inflammatory pathways that were not captured by CRP. Future research could examine whether markers such as IL-6, particularly at later developmental stages, are elevated in relation to these sustained cognitive stress processes.

### Neurobiological Disparities in Depersonalisation and Derealisation

The finding that IL-6 relates to DP and CRP relates to DR supports the idea of distinct neurobiological underpinnings for each symptom. For example, DP has been linked to frontal cortex hyperactivation, while DR may involve disruptions in temporal-parietal regions (Murphy, 2023). Although this study did not use neuroimaging, previous research shows inflammation can alter brain connectivity, including across the default mode network (DMN) and salience network in individuals with dissociation (Schmitz et al., 2025). We speculate that distinct inflammatory markers may preferentially influence different brain regions implicated in DP and DR. For instance, early-life IL-6 exposure could influence DMN development, impairing self-awareness and identity processing (such as self-concept clarity, Lassri et al., 2023), which could result in DP later. This is supported by a study that found reduced functional connectivity between the extrastriate body area (EBA) and the DMN predicted higher DP in depression (Paul et al., 2019), suggesting a potential link between neuroinflammation, DMN-EBA connectivity, and dissociative symptoms.

### Allostatic Load as a new framework for understanding DPDR

Beyond absolute levels of inflammatory markers, attention is shifting to the concept of biological instability. This is often conceptualized through the framework of allostatic load (AL), which is defined as “the cost of chronic exposure to a fluctuating or heightened neural or neuroendocrine response resulting from repeated or chronic environmental challenge” (McEwan & Stellar, 1993).

Measuring AL is critical due to its association with worse psychological outcomes, including increased risks of depression, anxiety, and suicide (Gou et al., 2025). While DPDR are traditionally seen as dissociative symptoms, they may also reflect prolonged physiological stress responses, making them particularly relevant to an AL framework. This study is the first to explore whether inflammatory dysregulation, a core feature of allostatic overload, predicts the persistence of DPDR over time.

Individuals experiencing DPDR often have a history of chronic stress, such as ACEs (Thomson & Jaque, 2018; Vonderlin et al., 2018; King et al., 2020; Quiñones, 2022), which are also common in those with dissociative PTSD (Schalinski et al., 2016). Dissociative disorders with childhood maltreatment are linked to smaller amygdalae and hippocampal volumes (Vermetten et al., 2006), brain areas highly associated with chronic stress and AL (Danese & McEwen, 2012). Thus, the presence of DPDR, its relationship with low-grade systemic inflammation, and evidence of immune dysregulation in DPDR, suggest these symptoms could be a manifestation of “allostatic overload” (McEwen, 2004). Importantly, stress may become chronic not only due to external adversity but also because it becomes self-sustaining once internalised. DPDR itself can act as a persistent stressor (e.g., the cognitive-behavioural conceptualisation of DPDR; Hunter et al., 2003), perpetuating physiological dysregulation and contributing to a cycle of allostatic overload.

### Strengths and Limitations

This study offers several key strengths. First, its large, population-based longitudinal dataset (IL-6 = 2,388; CRP = 3,774) enabled robust tracking of inflammatory markers and dissociative symptoms across developmental stages. This population sample provides a more representative view of DPDR than smaller clinical studies, which is crucial given that most cases of DDD go undiagnosed for up to 12 years (Baker et al., 2003; Michal et al., 2016).

Second, although DP and DR are classified together and frequently co-occur, they are phenomenologically distinct: DP involves alterations in self-experience, while DR reflects changes in the perception of the external world. Yet they share core features, such as a subjective sense of detachment from reality. Despite their overlap in clinical presentation and shared diagnostic category, our findings suggest they may be underpinned by different biological mechanisms. Recognising these distinctions is important for advancing understanding of DPDR and may help inform more targeted research and clinical strategies for diagnosis, prognosis, and treatment.

Finally, our findings’ grounding in the AL framework enhances their clinical relevance. This framework explains how low-grade systemic inflammation, reflecting chronic stress, may contribute to DPDR persistence. Positioning DPDR as a biologically embedded stress outcome highlights the potential for physiological interventions alongside traditional psychological therapies, improving future identification and treatment strategies.

Despite its strengths, this study has several limitations. First, IL-6 was measured only at age 9 years, limiting insights into its longitudinal changes and sustained versus transient inflammation. Second, DPDR were assessed using binary outcomes based on single items. While consistent, this did not capture symptom severity or frequency, constraining our ability to assess dose-response relationships or subthreshold variation. Finally, while associations were found, this study cannot determine causality; it remains unclear if inflammation directly contributes to symptom development, or if dissociation/upstream factors like chronic stress drive inflammatory changes.

### Clinical Implications and Future Directions

Research should prioritise investigating inflammatory markers or AL indices in relation to DPDR. This study represents a significant advancement, offering potential for symptom management. While trauma-focused treatments are first-line, immune-modulating strategies may be beneficial, especially for patients with elevated IL-6 or CRP fluctuations. Interventions targeting systemic inflammation (Gaesser et al., 2012; Klooster et al., 2020), like omega-3 supplementation, probiotics, exercise, and cytokine-targeting therapies, as well as and psychological interventions (O’Toole et al., 2018) could be explored as adjunctive treatments for DPDR in individuals with inflammatory dysregulation.

A key limitation of this study is that within ALSPAC, DP and DR were only measured as binary outcomes (present vs. absent), which restricted our ability to examine symptom severity or frequency. Future research should use dimensional assessments to capture the full spectrum of DPDR experiences and enable more precise analysis of their relationship with inflammatory markers. Additionally, this would allow for a more thorough investigation of inflammatory markers and symptom specificity, as well as observe a potential dose-response relationship between inflammatory markers and DPDR severity and/or frequency. Additionally, future research should examine the impact of anti-inflammatory treatments on DPDR prevalence. Drawing on evidence that CRP levels can guide antidepressant selection in depression (e.g., better outcomes with monotherapy for CRP <1 mg/L vs. bupropion-SSRI for >1 mg/L; Jha et al., 2017), CRP-stratified treatment approaches could be explored for DPDR. This might involve targeting norepinephrine/dopamine systems or anti-inflammatory agents for those with elevated/unstable CRP, versus traditional SSRIs or non-pharmacological approaches for those with low, stable CRP. Additionally, future research could explore how inflammatory processes may contribute to or modulate neural activity in DPDR. For instance, EEG has revealed altered perceptual and emotional processing in individuals with high DPDR (Gillmeister et al., 2024), and inflammatory markers could provide insight into the physiological mechanisms driving these neural differences.

## Conclusion

This study provides the first longitudinal evidence linking systemic inflammation (as measured via IL-6 and CRP) to the persistence of DP and DR, reinforcing the potential for biological mechanisms underlying these experiences. Drawing on a large, population-based sample, these results are likely to generalise beyond clinical settings, underscoring the relevance of immune system functioning for understanding chronic dissociative symptoms in the general population. By identifying biological correlates of DPDR, future research can explore inflammation as a possible target for intervention in disorders of self and perception.

## Data Availability

Access to ALSPAC data is granted via a detailed application process, ensuring participant privacy and ethical oversight; further information is available on the ALSPAC website (https://www.bristol.ac.uk/alspac/).

## Acknowledgements

We gratefully acknowledge Professor Paul Clarke for his expert statistical consultation and guidance on this work. The UK Medical Research Council and Wellcome (Grant ref: 217065/Z/19/Z) and the University of Bristol provide core support for ALSPAC. This publication is the work of the authors and Evelyn Dilkes will serve as the guarantor for the contents of this paper. This research was conducted as part of a PhD funded by the Economic and Social Research Council and the Biotechnology and Biological Sciences Research Council (Grant ref: ES/T00200X/1). We are extremely grateful to all the families who took part in the ALSPAC, the midwives for their help in recruiting them, and the entire ALSPAC team, which includes interviewers, technicians, clerical workers, research scientists, volunteers, managers, receptionists, and nurses.

