## Supplementary Information for "Interleukin-6 predicts depersonalisation and C-reactive protein predicts derealisation: A longitudinal study using the Avon Longitudinal Study of Parents and Children dataset"

**Supplementary Info**

1. *Cohort Description and Sample Sizes*

This study used mother, partner and child data from ALSPAC (Harris et al., 2009; Boyd et al., 2013; Fraser et al., 2013; Northstone et al., 2019; 2023). Pregnant women residing in Avon, UK with expected dates of delivery between 1^st^ April 1991 and 31^st^ December 1992 were invited to take part in the study. Of the initial enrolled 14,541 pregnancies, there was a total of 14,676 foetuses, resulting in 14,062 live births and 13,988 children who were alive at one year of age. When the oldest children were approximately 7 years of age, an attempt was made to bolster the initial sample with eligible cases who had failed to join the study originally. As a result, data are available for 15,447 pregnancies and 14,901 children alive at 1 year of age for all variables collected (and potentially abstracted from obstetric notes) from the age of 7 onwards. Parents completed annual postal questionnaires reporting on their child’s health and development from birth. From age 7, children also attended clinic-based assessments involving structured interviews and physical examinations. Informed consent for the use of data collected via questionnaires and clinics was obtained from participants following the recommendations of the ALSPAC Ethics and Law Committee at the time. Ethical approval was obtained from the ALSPAC Law and Ethics committee and the local research ethics committees. Study data were collected and managed using REDCap electronic data capture tools hosted at the University of Bristol (Harris et al., 2009). REDCap (Research Electronic Data Capture) is a secure, web-based software platform designed to support data capture for research studies.

2. *Additional Analyses: Cross-Sectional and Longitudinal Associations Between CRP at Age 9 and DP/DR*

This supplementary analysis presents two sets of results: (1) cross-sectional associations between CRP at age 9 and symptoms of depersonalisation (DP) and derealisation (DR) at age 12; and (2) longitudinal associations between CRP at age 9 and DP and DR symptoms at ages 17 and 24, with estimates interpreted relative to baseline symptom levels at age 12.

These analyses were conducted for consistency with the IL-6 models, which only included a single inflammatory marker (measured at age 9) and examined long-term effects on later DP and DR outcomes. While CRP was available at multiple time points, this analysis isolates the predictive value of CRP at age 9 to allow direct comparison with the IL-6 findings. As this specific approach was not central to the main hypothesis and is not discussed in the primary results, it is presented here for completeness.

CRP at 9 years was not significantly associated with DR at 12 years (aOR = 1.07, 95% CI: 0.93 - 1.21, p = .341). Further, in relation to DR scores at age 12, CRP did not significantly predict increased or decreased odds of experiencing DP at age 17 (aOR = 0.71, 95% CI: 0.43 – 1.17, p = .179). However, higher CRP levels at age 9 were associated with a decreased likelihood of DR symptoms at age 24 in comparison to baseline (aOR = 0.27, 95% CI: 0.16 – 0.44, p < .001), indicating a 73% decrease in the likelihood of DR symptoms.

*Supplementary Table 1: Unadjusted and adjusted OR for the association between log-transformed CRP at age 9 and DP and DR at ages 12, 17, and 24*

|  | Unadjusted Model | | | Adjusted Model | | |
| --- | --- | --- | --- | --- | --- | --- |
| **Age** | ***OR*** | ***CI*** | ***P*** | ***aOR*** | ***CI*** | ***P*** |
| **Depersonalisation** | | | |  |  |  |
| 12 | 0.99 | 0.89 - 1.11 | .846 | 0.94 | 0.83 - 1.05 | .267 |
| 17 | 0.93 | 0.52 – 1.64 | .791 | 0.90 | 0.49 – 1.65 | .727 |
| 24 | 0.83 | 0.47 – 1.45 | .505 | 0.74 | 0.41 – 1.35 | .325 |
| **Derealisation** | | |  |  |  |  |
| 12 | 1.10 | 0.97 - 1.23 | .127 | 1.07 | 0.93 – 1.21 | .341 |
| 17 | 0.77 | 0.49 – 1.20 | .247 | 0.71 | 0.43 – 1.17 | .179 |
| 24 | 0.26 | 0.16 – 0.41 | **< .001** | 0.27 | 0.16 – 0.44 | **< .001** |
